# Performance of EasyBreath^®^ Decathlon Snorkeling mask for Delivering Continuous Positive Airway Pressure

**DOI:** 10.1101/2020.10.28.20221317

**Authors:** Alberto Noto, Claudia Crimi, Andrea Cortegiani, Massimiliano Giardina, Filippo Benedetto, Pietro Princi, Annalisa Carlucci, Lorenzo Appendini, Cesare Gregoretti

## Abstract

**Background:** During the COVID-19 pandemic, the need for noninvasive respiratory support devices has dramatically increased, sometimes exceeding hospital capacity. The full-face Decathlon snorkeling mask, EasyBreath^®^ (EB^®^ mask), has been adapted to deliver continuous positive airway pressure (CPAP) as an emergency respiratory interface. We aimed to assess the performance of this modified EB^®^ mask.

**Methods:** CPAP set at 5, 10, and 15 cmH_2_O was delivered to 10 healthy volunteers with a high-flow system generator set at 40, 80, and 120 L min^-1^ and with a turbine-driven ventilator during both spontaneous and loaded (resistor) breathing. Inspiratory CO_2_ partial pressure (PiCO_2_), pressure inside the mask, breathing pattern and electrical activity of the diaphragm (EAdi) were measured at all combinations of CPAP/flows delivered, with and without the resistor.

**Results:** Using the high-flow generator set at 40 L min^-1^, the PiCO_2_ significantly increased and the system was unable to maintain the target CPAP of 10 and 15 cmH_2_O and a stable pressure within the respiratory cycle; conversely, the turbine-driven ventilator did. EAdi significantly increased with flow rates of 40 and 80 L min^-1^ but not at 120 L min^-1^ and with the turbine-driven ventilator.

**Conclusions:** EB^®^ mask can be safely used to deliver CPAP only under strict constraints, using either a high-flow generator at a flow rate greater than 80 L min^-1^, or a high-performance turbine-driven ventilator.

## INTRODUCTION

Hospitals and physicians worldwide are facing a new health emergency as a consequence of the coronavirus diseases 2019 (COVID-19) pandemic ^1^ that has spread enormously worldwide ^2^, placing an extraordinary demand on the health-care systems. Approximately 30% of hospitalized patients with COVID-19 develop acute hypoxemic respiratory failure (AHRF) requiring oxygen and noninvasive respiratory support (NRS). About 5% of them require invasive mechanical ventilation (IMV) and intensive care unit (ICU) admission ^3-5^.

Healthcare organizations have proactively implemented several strategies to supply shortages of equipment as demand surges, rationing life-saving treatments such as using shared mechanical ventilation ^6^, establishing ventilator lottery or ventilator triage policies ^7,8^, and increasing the use of NRS ^9^. To address the high demand for NRS equipment exceeding the capacity, Food and Drug Administration issued an emergency policy allowing the use of home respiratory devices, such as Continuous Positive Airway Pressure (CPAP), as an alternative to life-saving ventilators ^10^.

The need to ensure the availability of the highest possible number of CPAP devices also led a group of engineers from northern Italy to build an “emergency ventilator mask”, converting full face snorkeling masks into interfaces that can be used to apply CPAP therapy ^11^. Nevertheless, innovative and potentially life-saving products not previously tested and approved for clinical use can cause serious adverse effects.

Thus, the present study aimed to assess the technical performance of a modified commercial surface snorkeling mask that has been used in COVID-19 patients in delivering CPAP. The aim of this study was to evaluate the stability of pressure generated and the amount of carbon dioxide (CO_2_) rebreathing inside the mask during spontaneously and resistive loaded breathing in healthy volunteers.

## RESULTS

Ten healthy volunteers (eight males and two females), with age comprised between 27 and 45 years old and a mean body mass index (BMI) of 24 kg/m^2^ completed the study. None of the subjects complained about discomfort or fogging of the interface.

The FiO_2_ delivered during CPAP with the flow generator was between 31% and 34%, depending on the flow/PEEP combination. During CPAP delivered with the turbine-driven ventilator, the FiO_2_ ranged from 30 to 35%.

The PiCO_2_ ranged from 0 to 7 mmHg, depending on the flow/CPAP level combination used (p<0.01, ANOVA for repeated measurements), as shown in Table 1. The highest PiCO_2_ was recorded at 40 L min^-1^ of fresh gas flow with CPAP 15 cmH_2_0 and it decreased progressively to 0 (undetectable) as the fresh gas flow increased up to 120 L min^-1^ or using the turbine-driven ventilator, independently of the CPAP level.

**Table 1.**
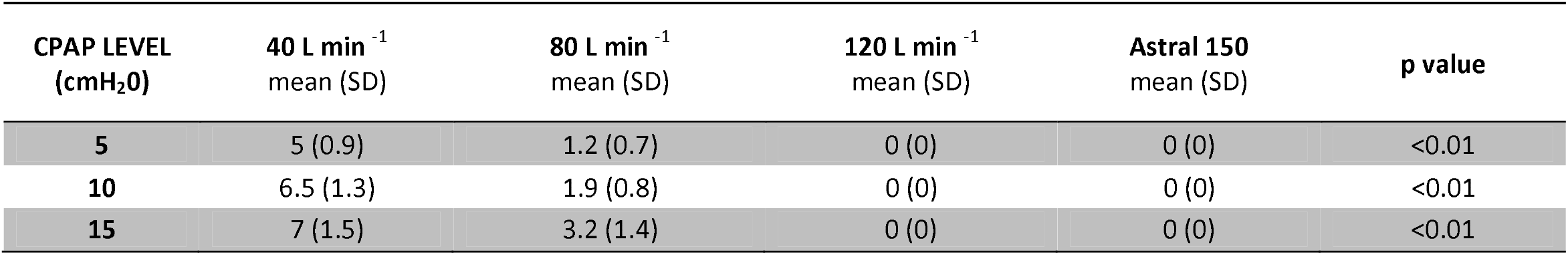
Inspiratory CO_2_ partial pressure at different flow rate and PEEP level. Abbreviations: CPAP, continuous positive airway pressure; SD, standard deviation; PEEP, Positive End Expiratory Pressure; Astral 150: turbine-driven ventilator. p value ANOVA for repeated measures (same CPAP level with different flow).

| CPAP LEVEL<br>(cmH<sub>2</sub>O) | 40 L min<sup>-1</sup><br>mean (SD) | 80 L min<sup>-1</sup><br>mean (SD) | 120 L min<sup>-1</sup><br>mean (SD) | Astral 150<br>mean (SD) | p value |
| --- | --- | --- | --- | --- | --- |
| 5 | 5 (0.9) | 1.2 (0.7) | 0 (0) | 0 (0) | <0.01 |
| 10 | 6.5 (1.3) | 1.9 (0.8) | 0 (0) | 0 (0) | <0.01 |
| 15 | 7 (1.5) | 3.2 (1.4) | 0 (0) | 0 (0) | <0.01 |

The pressures inside the mask at different flows/PEEP are shown in Table 2 and Figure 3. Target CPAP was not achieved when the oxygen-driven flow generator delivered 40 L min^-1^. Only the highest flow tested (120 L min^-1^) allowed the achievement of all the CPAP levels tested (5,10,15 cmH_2_O), even if some over-treatment was obtained at CPAP 5 and 10 cmH_2_O (+ 119% and + 35%, respectively). By contrast, the turbine-driven ventilator allowed the maintenance of target CPAP levels throughout the protocol.

**Table 2.**
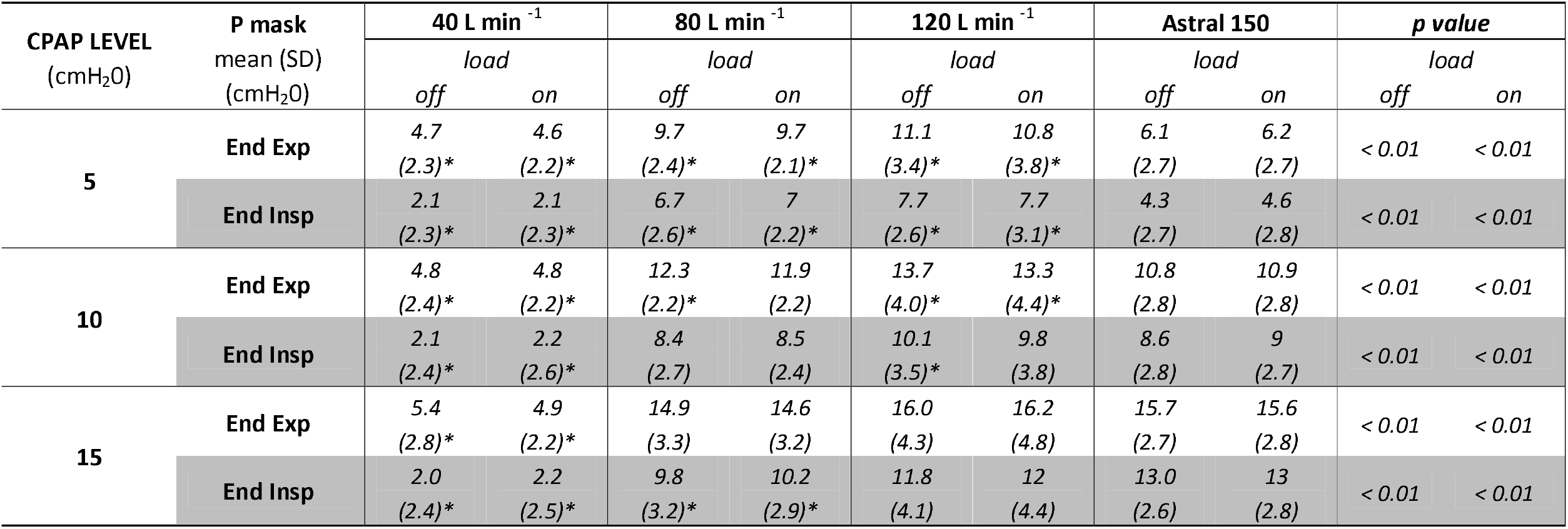
Pressure recorded inside the mask (P mask) during different experimental settings. Abbreviations: CPAP, continuous positive airway pressure; SD, standard deviation; End Exp, end-expiratory; End Insp, end-inspiratory; load, tube connector with inner diameter of 5 mm; Astral 150: turbine-driven ventilator. *: Significant difference compared to Astral 150 (Within-Subjects Contrast); p value: repeated measures ANOVA (same CPAP level at different flow).

Table 2 also shows actual CPAP partitioned between the inspiratory and expiratory phase, as referenced to target CPAP. Only the turbine ventilator succeeded in keeping the actual CPAP level close to its target value both at end-expiration and at end-inspiration for all the CPAP levels tested, the pressure difference between end-inspiration and end-expiration being maintained within 2 cmH_2_O. Statistically and clinically worse was the performance of the high-flow generator in this task. As a matter of fact, at a flow of 40 L min^-1^, neither end-inspiratory nor end-expiratory pressures reached the target CPAP. In contrast, at 80 and 120 L min^-1^, the actual pressure was above its target level at end-expiration and below it at end-inspiration, with the difference between these two conditions being well above 2 cmH_2_O, Table 2. Similar results were obtained for both loaded and unloaded breathing, Table 2.

The respiratory pattern (flows, volume, and respiratory timing) and the EAdi recorded at baseline and at different experimental conditions, with and without the use of the resistor, are shown in Table S1 and Figure 4. In comparison to baseline, minute ventilation remained stable with the CPAP mask at any level of pressure applied during unloaded breathing. By contrast, it significantly decreased during loaded breathing because of a significant reduction of tidal volume. Respiratory rate remained constant throughout the protocol (Table S1).

Diaphragmatic electrical activity increased with any CPAP level compared to baseline, either in terms of EAdi Insp_AUC_ and EAdi/V_T_ (Fig. 4). During loaded breathing, EAdi increased by a similar amount in all the conditions tested.

## DISCUSSION

The present study showed that the use of a modified EB^®^ mask coupled with a high-flow generator: 1) induces substantial CO_2_ rebreathing when set at a flow rate below 80 L min^-1^, 2) fails to achieve CPAP levels higher than 5 cmH_2_O if set at a flow rate of 40 L min^-1^ and 3) increases diaphragm’s electrical activation (and, hence, diaphragmatic energy expenditure) due to pressure instability along the respiratory cycle. These adverse effects disappear using high flows flushing the CPAP circuit (> 80 L min^-1^) or a high-performance turbine-driven ventilator set in CPAP mode.

The use of repurposed devices to fend off the shortage of ventilators during the current COVID-19 pandemic, without prior evidence of efficacy, is an issue of increasing concern ^12^. Manufacturers are allowed to market many medical devices without the need to provide proof of their effectiveness, especially in urgent unusual circumstances like the COVID-19 pandemic ^13^. Indeed, clinicians may be forced to adopt unregistered interfaces, such as the popular Decathlon EB^®^ mask, to deliver ventilatory support to patients with little or no prior tests of performance. To the best of our knowledge, this is the first study that fills this knowledge gap after the introduction of the modified Decathlon EB^®^ mask for CPAP delivery.

The use of CPAP in acute hypoxemic respiratory failure has the principal aim to improve gas-exchange increasing the end-expiratory lung volume (EELV) and avoiding at the same time the inspiratory elastic threshold-load imposed by the increased EELV ^14^.

This task has been effectively accomplished by non-invasively applied CPAP ^15,16^. However, some interfaces can provoke CO_2_ rebreathing if misused, thus impairing treatment effectiveness ^17^.

### CO_2_ rebreathing

EB^®^ mask has a wide dead-space (about 800-900 ml), and thus, given a fixed CO_2_ production, it requires high flows to wash-out the exhaled CO_2_ completely. In accordance with previous findings obtained with other high dead-space interfaces ^17^ only CPAP flow rate of 120 L min^-1^ or flows guaranteed by a high-performance turbine-driven ventilator completely flushed out CO_2_. Thus, in case of emergency use of the EB^®^ mask, the first caveat is to use very high flows of fresh gas (about fourfold the peak inspiratory flow) in the circuit, whatever its source.

### Effectiveness in achieving target CPAP

An ideal CPAP system, by definition, should provide a target positive airway pressure through the respiratory cycle to improve alveolar recruitment and gas exchange. This was not the case of the EB^®^ mask. Our data show that although the pressure inside the mask never dropped below zero during the inspiratory phase, it always remained far below the set CPAP level when using a fresh gas flow of 40 L min^-1^. On the other hand, during the expiratory phase, the pressure level recorded at the highest rate of fresh gas flow (120 L min^-1^) overcame the set CPAP level, at least at 5 and 10 cmH_2_O, potentially generating an undue increase in end-expiratory lung volume. Fresh gas flow of 80 L min^-1^ performed in the middle, overcoming the set pressure of 5 cmH_2_O and under-assisting at the target CPAP of 15 cmH_2_O.

Mechanical ventilators are the most sophisticated devices to administer CPAP. However, old devices performed worse than high flow generators in delivering stable CPAP levels, mainly because of the demand-valve technology ^18^. The newer generation of ventilators has complex flow algorithms that allow for a more aggressive flow modulation and delivery flows using single circuits with intentional leaks designed to stabilize airway pressure and lessen the imposed work of breathing ^15^. This was the case in our study when CPAP was delivered with the turbine-driven ventilator provided with an intentional leak circuit (EB^®^ mask + turbine-ventilator + single circuit + intentional leak), where we recorded a stable pressure at any level of target CPAP tested.

### Diaphragmatic activity and effort

An ideal CPAP system should provide a constant positive airway pressure throughout the respiratory cycle to avoid negative effects on the work of breathing ^19,20^ since large pressure swings around the set PEEP level are associated with increased respiratory efforts ^21^. Our study showed that end-inspiratory to end-expiratory pressure swing was clinically relevant (above 3 cmH_2_0) at all set levels of CPAP and flows when using the high flow generator with a flow rate lower that 80 L min^-1^. We observed this phenomenon notwithstanding the presence of a pressure stabilizer (reservoir bag) and a threshold PEEP valve in the CPAP circuit that should maintain the pressure stable whatever is the flow passing through it. Pressure swings occurring across target CPAP levels imply wasted patients’ respiratory effort spent against CPAP device, ineffective in producing flow and volume ^18^. As a matter of fact, EAdi inspiratory activity and, hence, diaphragmatic effort ^22^ increased with CPAP application compared to baseline in all the study conditions (with or without resistive loading) when high flow CPAP generator delivered flows of 40 and 80 L min^-1^. This result confirms previous data showing increased work of breathing during CPAP application ^23^, and suggesting a suboptimal performance of high flow CPAP generators ^24,25^.

EAdi increase can be due to CPAP system-induced increased workload, on a CO_2_ rebreathing-induced increase of respiratory drive, or a combination of both. The inspiratory EAdi-time product (EAdi InspAUC) increased by 45% during CPAP application and only 35% when it was normalized for V_T_, indicating that both the above-quoted mechanisms contributed to the overall increase of EAdi activity during EB^®^ mask CPAP delivery, as compared to spontaneous breathing. The increase of diaphragmatic electrical activity (and effort) disappeared during CPAP set at the highest flow tested (120 L min^-1^) and during CPAP delivered by the turbine ventilator indicating that these constraints should be considered in case of emergency use of the EB^®^ mask.

This study should be a warning sign against the indiscriminate use of tools to deliver noninvasive respiratory support without rigorous tests of performance.

## Strengths and limitations

The main strength of our study is the rigorous evaluation of several parameters of performance (i.e. PiCO_2,_ Pmask, EAdi) at three different levels of flow, recreating the setup used in clinics for noninvasive ventilatory support in COVID-19 patients.

The present study has several limitations. First, this was a proof-of-concept study performed in healthy volunteers; therefore, it may not reflect the dynamic nature of ventilation parameters during severe acute respiratory syndrome. However, simulating loaded breathing, we partially overcame this limitation. Furthermore, we did not compare the new device against a ‘gold standard’. Thus, up to now, it is only clear that the EB^®^ mask works properly only under strictly controlled conditions. Still, it is impossible to state whether it is inferior/superior to reference interfaces. Finally, the PiCO_2_ was stable during the conditions tested, but the recordings lasted only 2 minutes each. Therefore, we cannot ensure that these values would have remained stable for a longer period.

## CONCLUSION

The modified Decathlon’s EB^®^ mask should be used for emergency use to deliver CPAP only under strict constraints, using a high-flow generator at a flow rate greater than 80 L min^-1^, or with a high-performance turbine-driven ventilator.

## METHODS

### Study Design

We designed a physiological study involving ten healthy volunteers recruited among physicians at the Department of Anesthesia and Critical Care, Policlinico “G. Martino,” University of Messina, Italy, in June 2020. The study was approved by the Ethics committee of Policlinico G. Martino, Messina, Italy (32-30 27/05/2020). All the study participants provided written informed consent.

### Experimental set-up and Study devices

The experimental set-up is illustrated in Figure 1. We used a modified Decathlon’s EasyBreath^®^ (EB^®^ mask) surface snorkeling mask (Decathlon, Villeneuve-d’Ascq, France) as a full-face mask to deliver CPAP. The original valve set on the chin was modified to be permanently closed, allowing all exhaled air to be evacuated through the two lateral channels of the mask to the top of the snorkel ^26^. Unlike conventional interfaces used in clinical practice, this mask has two different compartments inside; the upper one for the eyes, dedicated to the vision, and the lower one for the mouth and nose, dedicated to the breathing. The original EB^®^ mask snorkel was replaced with a 3D printed adaptor, the Charlotte valve, patented by ISINNOVA (Brescia, Italy), to connect the mask to an oxygen-driven flow generator. This valve has an inlet line connected to the oxygen and an outlet line connected to a threshold resistor [Positive End Expiratory Pressure (PEEP) valve], as shown in Figure S1. The internal volume was measured by filling the mask with water when applied to the volunteers’ face, as previously described ^27^, and amounted to 880±11 ml.

**Figure 1:**
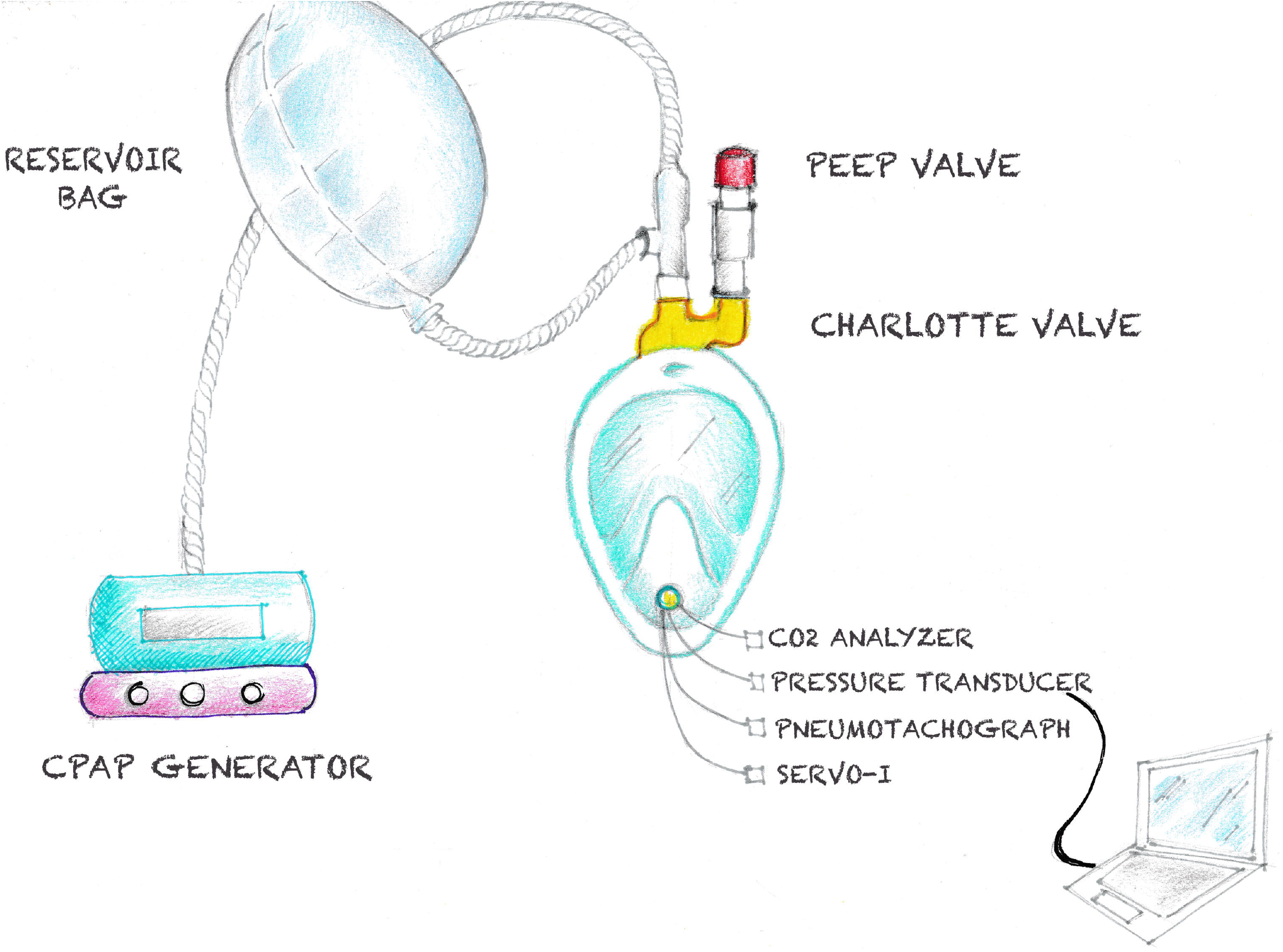
Experimental setup. Abbreviations: CPAP, continuous positive airway pressure; PEEP, Positive End Expiration Pressure.

To allow measurements, the experimental setting included the following items (figure 1):

1. a pneumotachograph (V.T. mobile; Fluke Biomedical, Everett, WA, USA) to measure ventilatory airflow, connected with a serial interface to a laptop for recording (V.T. for Windows Software version 2.01.07, Fluke Biomedical, Everett, WA, USA);
2. a side-stream capnometer (CO2SMO plus; Novametrix Medical System, Wallingford, Connecticut, USA) to measure inspiratory carbon dioxide tension (PiCO_2_);
3. a dedicated nasogastric tube with a built-in multiple array esophageal electrode (Edi catheter) to monitor diaphragm activity ^14^. It was connected to a Servo-I ventilator (Getinge, Solna, Sweden), equipped with Neurally Adjusted Ventilatory Assist (NAVA) (Getinge, Solna, Sweden), linked through a serial interface to a laptop with dedicated software (Servo-tracker version 4.2, Getinge, Solna, Sweden).
4. a pressure transducer (coupled to the V.T. mobile, through an auxiliary port) to measure the pressure inside the mask (Pmask) connected to the laptop for recording.

All the instruments underwent testing and calibration according to the manufacturers’ specifications before performing all the measurements.

### Measurements and recordings

Based on the experimental set-up, the following variables were measured: 1) PiCO_2_; 2) Minute ventilation; 3) Pmask; 4) electrical diaphragm activity (EAdi).

Each volunteer was studied placed comfortably in a semi-recumbent position and performed all the tests on the same day in random order. Volunteers were first allowed to adapt to CPAP breathing before the recordings.

At baseline, two, 120 seconds each, recordings of breathing pattern and EAdi were carried out without the mask. The first one during quiet unloaded breathing, and the second while breathing through an inline resistor (connected to the mouthpiece) made with an endotracheal tube connector with an inner diameter of 5 mm, to simulate an increase of respiratory load.

After tightly fitting to the volunteers’ face the modified EB^®^ mask ^11^, leaks were minimized by sealing the mask chin valve around all measurement lines. Two sets of measurements were performed while delivering CPAP with two different systems:

1. a CPAP flow generator (Whisperflow 2, Respironics), connected to the 3D printed Charlotte valve ^11^ attached to the mask. A reservoir bag inserted in the inspiratory line was used to minimize airway pressure variations. An adjustable PEEP valve (Ambu, Denmark) added to the Charlotte valve’s expiratory port provided the PEEP level. PiCO_2_, Pmask, flow lines, and the Edi catheter were inserted into the mask through the sealed chin valve (Figure 1). CPAP was set at three different flow rates (40, 80, 120 L min^-1^) and for each flow, three different PEEP levels were employed (5, 10, 15 cmH_2_O), with and without the resistor in place *(load)*;
2. a turbine-driven home care mechanical ventilator (Astral 150, Resmed, San Diego, USA) set in CPAP mode at three different PEEP levels: 5, 10, 15 cmH_2_0. CPAP was administered with a single limb vented configuration with an intentional leak (a tube connector with an inner diameter of 5.5 mm) placed on the Charlotte valve’s expiratory port. Low-pressure oxygen was provided to the turbine-driven ventilator through a dedicated port in order to obtain the same FiO_2_ used to drive the flow generator.

A 120 seconds-trial was recorded for each session for off-line analysis. No instructions were given to the subjects regarding the breathing pattern to adopt.

### Data analysis

A Matlab (version 9.7.0.1190202, The MathWorks Inc., Natick, Massachusetts, USA) automatic procedure was developed to find maximal and minimum EAdi (Figure 2) and pressure for each breath. The EAdi-time product, measured as the EAdi curve area from the beginning to the end of an inspiratory cycle (EAdi Insp_AUC_), was calculated (Figure 2). The EAdi Insp_AUC_ was then normalized for the tidal volume (V_T_), (EAdi /V_T_). Average end-expiration and end-inspiration peak pressures were calculated for every recording session. The same was also done for the EAdi_max_, EAdi Insp_AUC,_ and EAdi /V_T_. For each recording session, the average delta pressure inside the mask at the end-expiration (ΔPmask Exp = Pmask end-expiratory – CPAP Level) and end-inspiration (ΔPmask Insp = Pmask end-inspiratory - CPAP Level) were computed.

**Figure 2:**
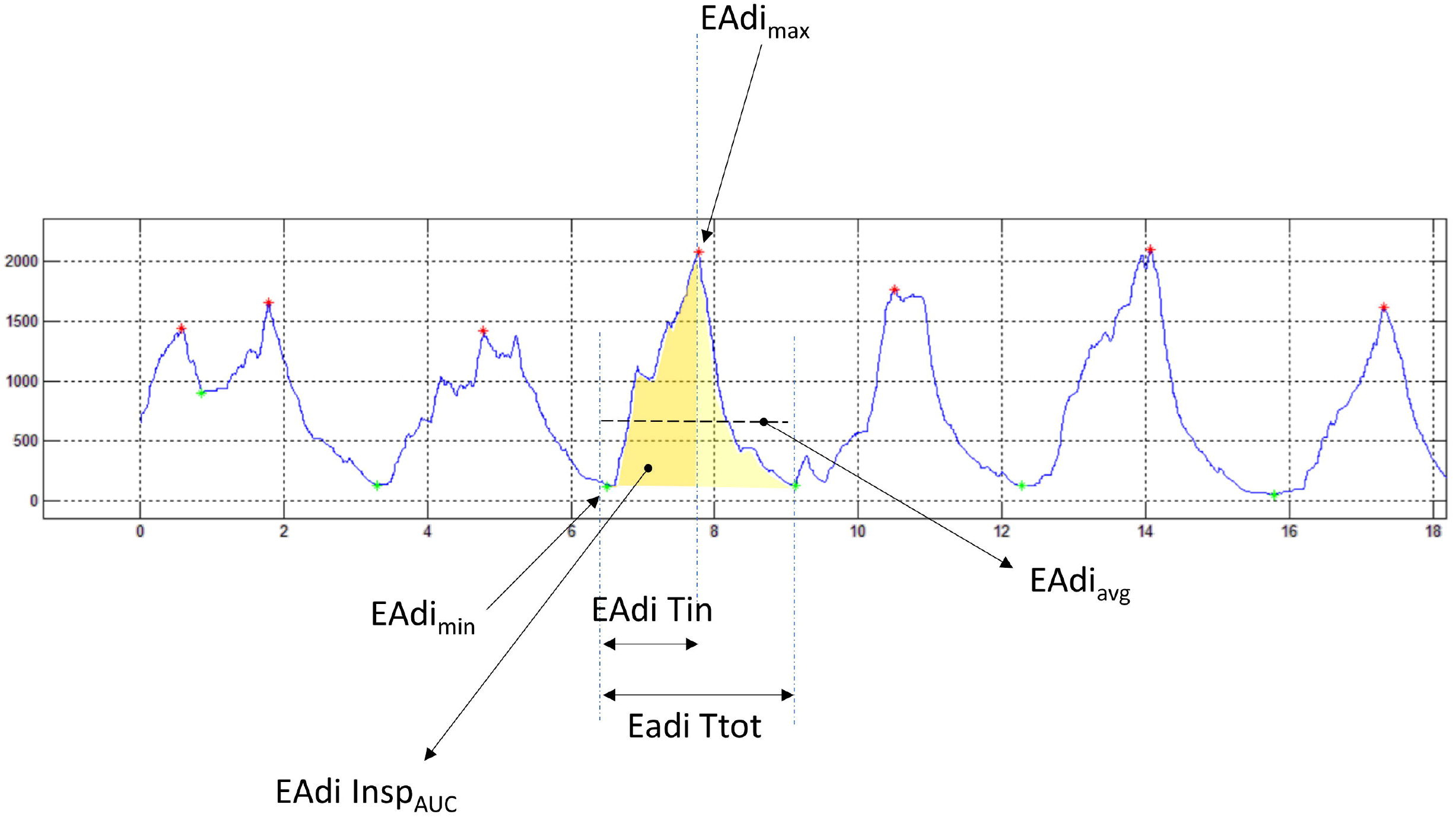
Matlab automatic EAdi analysis. Abbreviations: EAdi, electrical diaphragm activity; EAdi_max_, peak electrical diaphragm activity; EAdi_min_, electrical diaphragm activity; EAdi Insp_AUC_, electrical diaphragm activity area of the inspiratory phase; EAdi Tin, inspiratory time derived from electrical diaphragm activity; EAdi Ttot, total respiratory time derived from electrical diaphragm activity.

**Figure 3:**
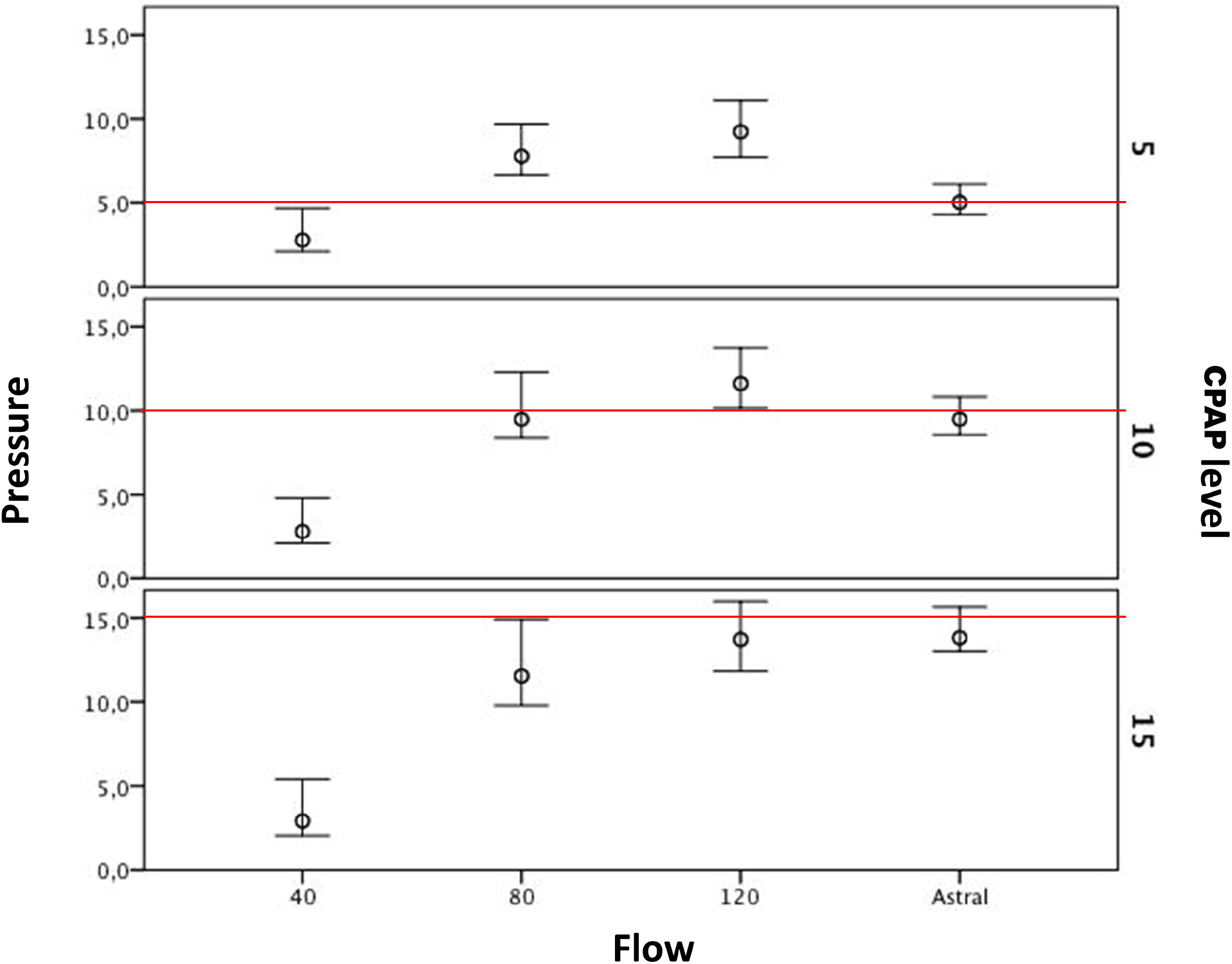
Pressure recorded inside the mask. Upper whiskers represent the mean pressure at end-expiration; Lower whiskers represent the mean pressure at end-inspiration; Circles represent the mean pressure; Redline represents the set CPAP level.

**Figure 4:**
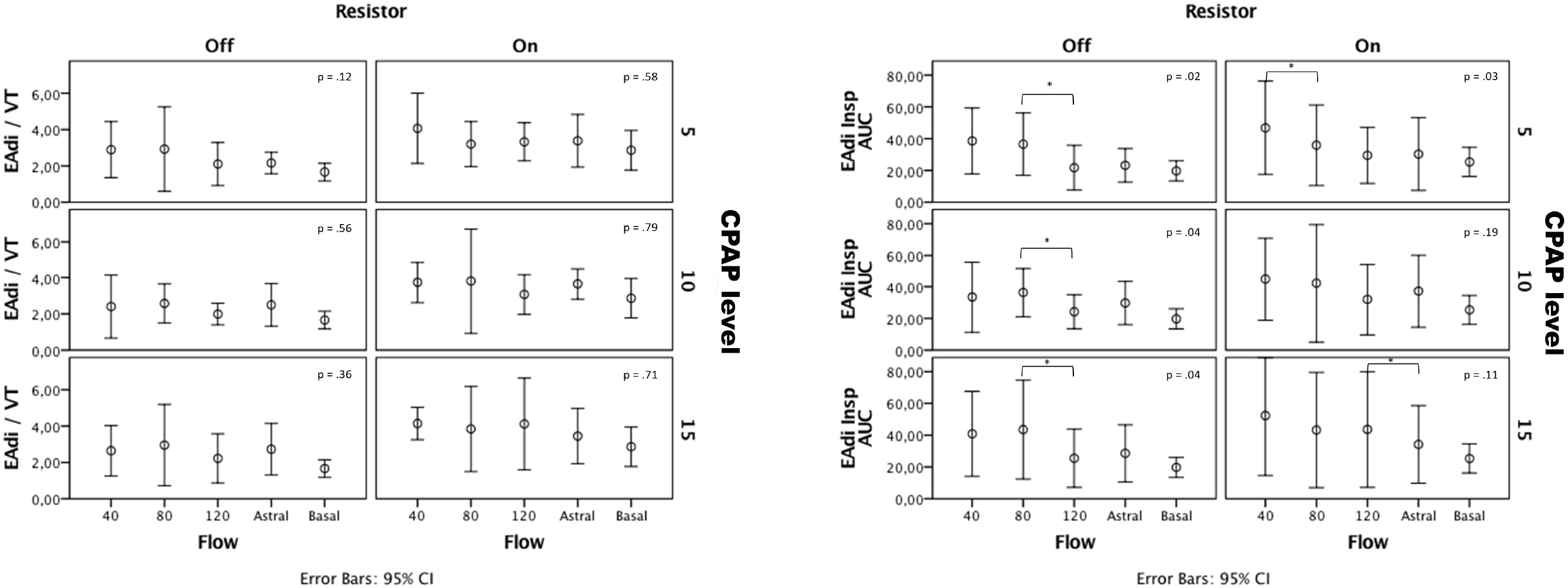
Electrical diaphragm effort. EAdi/V_T,_ electrical diaphragm activity area of the inspiratory phase normalized for the tidal volume (V_T_); EAdi Insp_AUC,_ electrical diaphragm activity area of the inspiratory phase.

The distribution of each continuous variable was initially tested with the Shapiro-Wilk test for normality. Results are expressed as mean ± SD when normally distributed, otherwise as median and interquartile range. Based on preliminary measurements, we estimated that 10 healthy subjects were necessary to detect, with a 0.8 power, a significant difference (α = .05) on airway pressure drop during inspiration (ΔPmask Insp). Comparisons of ΔPmask at different flows and for each CPAP level were assessed using the analysis of variance (ANOVA) for repeated measures. The statistical analysis was performed using (SPSS 24, IBM) software. A *p* value <.05 was considered significant.

## Data Availability

The data that support the findings of this study are openly available in repository data [figshare] at the link below.

https://doi.org/10.6084/m9.figshare.13151003

https://doi.org/10.6084/m9.figshare.13150577

## Acknowledgements

The authors express gratitude to Prof. G. Risitano for the 3D print of the Charlotte Valve and to M.L. Noto for assistance in the drawing.

## Authors’ contributions

A.N., C.G. and C.C, conceived and designed the study.

A.N., P.P., M.G. and F.B. collected the data.

A.N. and P.P., developed the data analysis tools.

A.N., P.P., A.C., A.Ca., L.A. performed the analysis.

A.N., C.C., L.A., C.G. wrote the manuscript.

All authors have critically revised the manuscript and approved the final version.

