## Supplementary material for "Performance of EasyBreath^®^ Decathlon Snorkeling mask for Delivering Continuous Positive Airway Pressure": Table S1

**Alberto Noto, MD, PhD**

Department of Human Pathology of the Adult and Evolutive Age “Gaetano Barresi”, Division of Anesthesia and Intensive Care, University of Messina, Policlinico “G. Martino”,

Via Consolare Valeria, 1

98100 Messina, Italy

**CONTENT**

Page 4	**Fig. S1:** Charlotte valve detailed view and inner pipes

Page 5	**Table S1:** Respiratory pattern and diaphragm activity

**Figure S1:** *Charlotte valve detailed view and inner pipes.*

a) valve top view; b) valve front view; c) inspiratory (blu) and expiratory (red) flow inside the mask; d) inspiratory (blu) and	expiratory (red) pipe inside the Charlotte valve.

|  | **Baseline** | | | **5 cmH_2_0** | | | **10 cmH_2_0** | | | **15 cmH_2_0** | | |
| --- | --- | --- | --- | --- | --- | --- | --- | --- | --- | --- | --- | --- |
|  | **load** | | | **load** | | | **load** | | | **load** | | |
|  | **Off** | **On** | **p** | **Off** | **On** | **p** | **Off** | **On** | **p** | **Off** | **On** | **p** |
| **VM, L min ^-1^** | *11(3.5)* | *7.2 (1.9)* | *<0.01* | *11.3 (4.3)* | *7.6 (2)* | *<0.01* | *11.8 (4.3)* | *7.6(2.2)* | *<0.01* | *11.5 (4.2)* | *8.0 (2.8)* | *<0.01* |
| **V_T_, ml^.^kg^-1^ IBW** | *11.9 (2.3)* | *9 (2.2)* | *<0.01* | *13.3 (5.1)* | *11.9 (5.3)* | *0.13* | *15.3 (5.1)* | *12.6 (5.8)* | *<0.01* | *15.6 (7)* | *13.3 (7.9)* | *<0.01* |
| **PIF, L min ^-1^** | *31.9 (9.8)* | *19.9 (3)* | *<0.01* | *32.4 (11.2)* | *19.3 (4.1)* | *<0.01* | *34 (13)* | *19.4 (5.2)* | *<0.01* | *32.4 (11.3)* | *19.9 (6.3)* | *<0.01* |
| **PEF, L min ^-1^** | *34.8 (9.1)* | *23.1 (5.8)* | *<0.01* | *38.9 (12.5)* | *23.6 (6.6)* | *<0.01* | *40.7 (13.1)* | *24.2 (6.7)* | *<0.01* | *41.3 (14.6)* | *25.5 (9.3)* | *<0.01* |
| **RR, breaths^.^min^-1^** | *12.7 (2.1)* | *11.1 (2.3)* | *<0.01* | *12.4 (5.3)* | *9.9 (3)* | *<0.01* | *10.9 (3.6)* | *9.5 (2.9)* | *0.03* | *11.3 (4.1)* | *10 (4.3)* | *0.07* |
| **Tin, s** | *3.1 (1.0)* | *3.3 (0.9)* | *0.02* | *3.87 (2.06)* | *4.20 (1.89)* | *0.13* | *4.00 (1.78)* | *4.60 (2.07)* | *<0.01* | *4.14 (1.75)* | *4.35 (1.93)* | *0.25* |
| **EAdi_max_, μV** | *37 (9)* | *33 (12* | *0.14* | *49.5 (22.8)* | *48.1 (26.9)* | *0.79* | *51.5 (19.7)* | *45.5 (20.8)* | *0.14* | *55.2 (25.8)* | *56.2 (30.9)* | *0.85* |
| **EAdi Insp_AUC,_ μV^2^** | *20 (8)* | *25 (10)* | *<0.01* | *27.5 (16.8)* | *34 (23.5)* | *0.06* | *28.4 (16)* | *36.0 (26.3)* | *0.02* | *31.3 (23.8)* | *39.3 (32.4)* | *0.04* |
| **EAdi / V_T_, μV^2 .^ml^-1^** | *1.66 (0.6)* | *2.86 (1.03)* | *<0.01* | *2.3 (1.2)* | *3.3 (1.3)* | *<0.01* | *2.2 (1)* | *3.4 (1.5)* | *<0.01* | *2.4 (1.4)* | *3.6 (1.7)* | *<0.01* |

**Table S1:** *Respiratory pattern and diaphragm activity*

Abbreviations: VM, volume minute; Vt, volume tidal; PIF, peak inspiratory flow; PEF, peak expiratory flow; RR, respiratory rate; Tin, inspiratory time; EAdi_max_, peak electrical diaphragm activity; EAdi Insp_AUC,_ electrical diaphragm activity area of the inspiratory phase; EAdi /V_T,_ electrical diaphragm activity area of the inspiratory phase normalized for the tidal volume (V_T_).
