## Supplementary figures and images for "Performance of EasyBreath^®^ Decathlon Snorkeling mask for Delivering Continuous Positive Airway Pressure"

### Figure S1

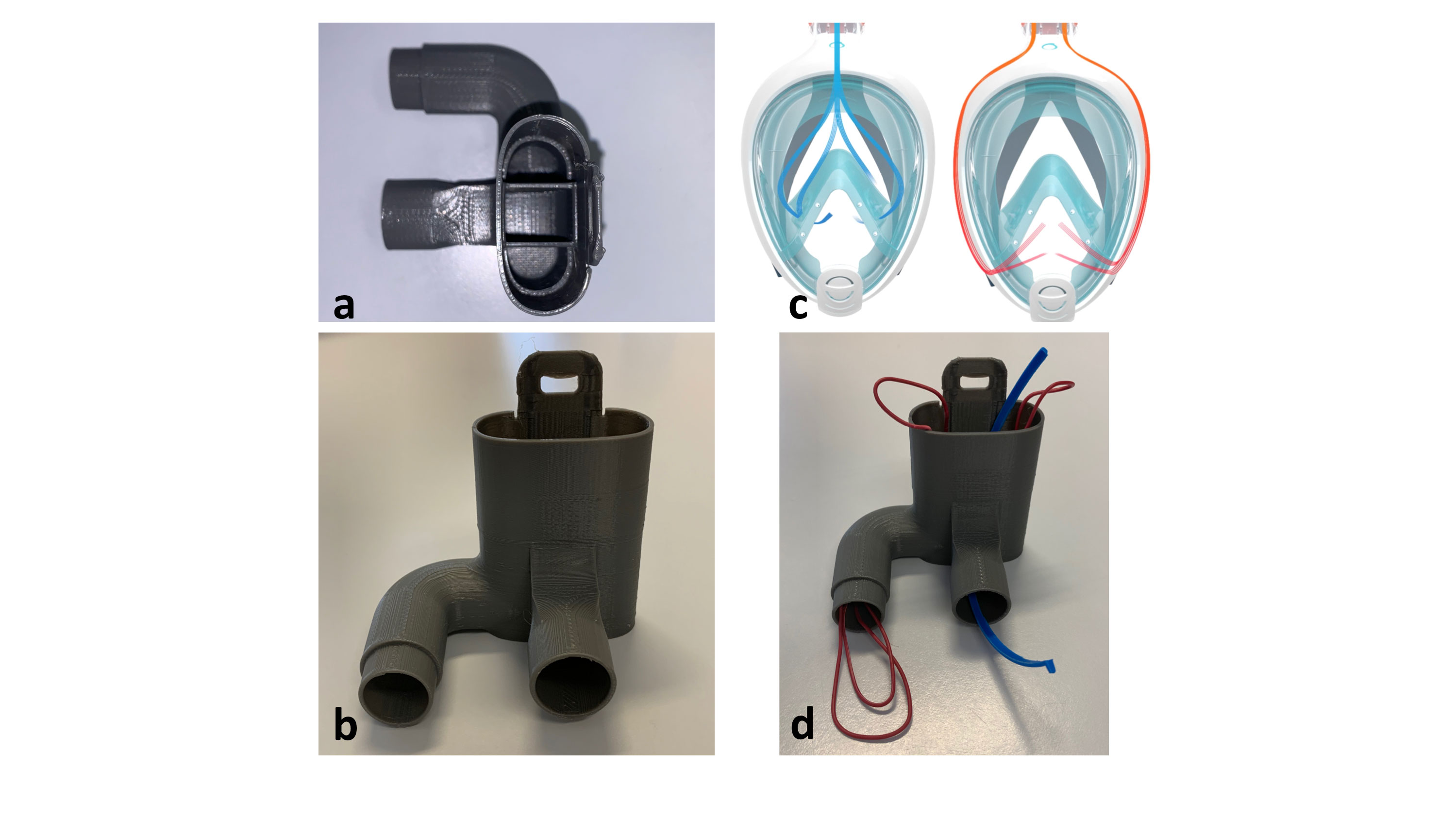
